# Standardisation in Acute Stroke Research: A Scoping Review of Upper Limb Assessments Against SRRR Benchmarks

**DOI:** 10.1101/2025.07.30.25332435

**Authors:** Milica Doric, Lisa Tedesco Triccas, Mingyao Xiong, Faye Tabone, Adrian L Knorz, Nicole Downar, Nick S Ward, Catharina Zich

## Abstract

**Background:** Stroke remains a leading global cause of long-term disability, with upper limb (UL) impairments affecting nearly two-thirds of survivors. The acute phase of stroke (1–7 days) is a critical window for understanding recovery mechanisms and evaluating interventions. To address inconsistencies in assessment and methodological approaches, the Stroke Recovery and Rehabilitation Roundtable (SRRR) has issued guidelines promoting standardisation across stroke research.

**Objective:** This scoping review examines how current acute stroke studies assessing UL sensorimotor capacity align with SRRR recommendations, particularly regarding measurement tools, follow-up protocols, and participant demographics.

**Methods:** Following the Joanna Briggs Institute and PRISMA-ScR guidelines, we systematically searched seven databases for English-language, primary research studies published between August 2017 and May 2025. Studies included adults with stroke who underwent UL assessment within the acute phase. Data were extracted on clinical, structural, and functional assessments, as well as follow-up timing and study demographics. Of 2,485 screened articles, 106 met the inclusion criteria.

**Results:** While global assessments (e.g., NIHSS) and impairment-level UL assessments (e.g., FMA-UE) were widely used, activity-level tools (e.g., ARAT) and sensory measures were underrepresented. Structural brain imaging was common, though often used only diagnostically, while functional brain imaging and multimodal approaches were rare. Follow-up timing varied, with limited long-term tracking. Demographic reporting was inconsistent, with underrepresentation of young adults and women.

**Conclusion:** Despite progress, significant gaps remain in the standardisation and comprehensiveness of UL assessment in acute stroke research. Future studies should better align with SRRR recommendations to improve data comparability and scientific rigour.

## 1. Introduction

The global prevalence of stroke has surpassed 101 million cases, with approximately 12.2 million individuals experiencing a first-ever stroke (Feigin et al., 2022; Writing Group Members et al., 2016). Stroke remains one of the leading contributors to disability-adjusted life years (Steinmetz et al., 2024). Notably, around two-thirds of stroke survivors go on to have upper limb (UL) sensorimotor impairments, often leading to long-term disability and reduced quality of life (Langhorne et al., 2011).

Although stroke is considered a chronic condition, it progresses rapidly during the hyper-acute (0-24 hours) and acute (1-7 days) stages (Bernhardt et al., 2017). Consequently, the acute stage represents a critical window for investigating the mechanisms underlying impairment, exploring therapeutic interventions, and identifying biomarkers predictive of recovery. However, research in the acute phase of stroke is fraught with practical challenges, particularly logistical barriers to timely patient recruitment (Broderick et al., 2023). Consequently, despite best efforts, studies in this domain are often limited by small sample sizes, significant heterogeneity in stroke pathophysiology, and inconsistent assessment methodologies - factors that collectively constrain the generalisability of findings. To address these issues and enhance the quality and impact of stroke research, the Stroke Recovery and Rehabilitation Roundtable (SRRR) has issued a series of recommendations aimed at promoting standardisation and fostering international collaboration in stroke recovery research.

Specifically, the SRRR emphasises the use of biologically meaningful post-stroke time windows - hyperacute (0-24h), acute (1-7days), early subacute (7 days – 3 months), late subacute (3-6 months), and chronic (>6 months) - to guide the timing of assessments and interventions (Bernhardt et al., 2017). The guidelines advocate for the systematic collection of baseline patient characteristics, including age, sex, time since stroke onset, stroke severity, and imaging confirmations, to enable more precise patient profiling (Kwakkel et al., 2017). The SRRR endorses a core set of validated clinical outcome measures to assess sensorimotor UL capacity (Kwakkel et al., 2017, 2019). Upper limb capacity is used as a unifying descriptor that encompasses the full spectrum of upper limb function, impairment, performance, and recovery, enabling consistent classification across diverse assessment tools and International Classification of Functioning, Disability and Health (ICF) outcome domains. Follow-up assessments are considered essential, with a 3-month evaluation identified as a critical timepoint for capturing meaningful recovery milestones (Kwakkel et al., 2017). The SRRR further highlights the role of biomarkers in elucidating recovery mechanisms. Structural and functional measures, particularly corticospinal tract (CST) integrity, are recognised for their utility in predicting sensorimotor outcomes, guiding interventions, and stratifying patients (Boyd et al., 2017). Achieving consensus on post-stroke stage definitions, as well as on the type and timing of assessments, is crucial for data pooling across studies, supporting robust meta-analyses, thereby fuelling new research insights.

Nearly one decade after the first SRRR meeting, it is timely to review the alignment of the research practices in acute stroke UL sensorimotor capacity with these recommendations and identify key gaps. We focused on the SRRR guidelines due to their specific relevance to stroke recovery and rehabilitation, particularly in standardizing outcome measures and research methodology. While other frameworks provide valuable insights, they are broader in scope; the SRRR offers a more targeted complement, aligning closely with the goals of our review. Specifically, this scoping review examines the demographic characteristics of participants in acute stroke studies on UL sensorimotor capacity. It also reviews the tools used to assess both global and UL-specific sensorimotor capacity, as well as changes in brain structure and function. In addition, it explores which assessment tools are commonly used in combination and the typical timing of follow-up assessments.

## 2. Methods

A focused scoping review, adhering to the proposed Joanna Briggs Institute (JBI) framework (Arksey & and O’Malley, 2005), was conducted to synthesise the literature, identify prevailing trends, and reveal existing gaps in acute stroke research. We followed the Preferred Reporting Items for Systematic Reviews and Meta-Analyses extension for Scoping Reviews (PRISMA-ScR) checklist (Tricco et al., 2018).

### 2.1. Search Strategy

Two researchers (CZ and MD) developed a formal search strategy based on combinations of key search terms related to “acute stroke,” “upper limb,” and “motor” (**Suppl. Table 1**). These terms were systematically applied across seven electronic databases – Embase (Ovid), MEDLINE (Ovid), PubMed, CINAHL (EBSCO), PsycINFO (Ovid), Google Scholar, and Web of Science - to identify candidate publications relevant to the study. The search was conducted on May 29, 2025, and restricted to articles available in English, those containing the specified keywords in the title and/or abstract, and publications between August 2017 and May 2025 (this time frame aligns with the emergence of the SRRR paper on agreed definitions for stroke (Bernhardt et al., 2017)). Grey literature databases were not included in this scoping review.

### 2.2. Eligibility Criteria

The eligibility criteria for this scoping review were informed by the JBI Population/Concept/Context (PCC) framework (Peters et al., 2020). To be included, studies needed to meet the following conditions:

#### Types of participants

Studies must involve stroke survivors, defined as individuals with a clinical diagnosis of stroke of any type, who have undergone assessments related to UL sensorimotor capacity in the acute stage post-stroke. Stroke survivors must be aged 18 years or over.

#### Concept

Our scoping review centers on determining how research studies quantify UL impairment in the acute stage based on the SRRR recommended clinical scales and measures of brain structure and function (Bernhardt et al., 2017; Boyd et al., 2017; Kwakkel et al., 2017, 2019). The inclusion of diverse methodologies provides a comprehensive overview of the diagnostic and evaluative tools currently in use within stroke research. Studies that do not specifically address UL function, such as those focusing on lower limb or other forms of capacity, were not included.

#### Context

Publications from all geographical locations and cultural contexts were included, provided that they were published in English.

#### Types of sources

Eligible sources comprise full-text, primary research studies, encompassing quantitative interventional and non-interventional studies (e.g., randomized and non-randomized controlled trials, pre-post studies, pilot studies, single-case experimental studies, case studies, case-control studies, cross-sectional studies, and cohort studies). Conversely, review articles, conference proceedings, protocols, clinical practice guidelines, perspectives, opinion pieces, and evaluations of new devices or measures were excluded, as they were considered inappropriate for addressing the objectives of this scoping review.

### 2.3. Selection of Sources of Evidence

All retrieved records (N = 2,485, Figure 1) were entered into EndNote 20 (Clarivate Analytics, PA, USA) referencing software. Duplicate removal was first performed automatically, followed by manual deduplication by MD to ensure accuracy (total number of duplicates = 1,171). Screening was conducted in multiple stages: title screening, abstract screening, and comprehensive full-text review. Title and abstract screening were conducted twice by trained independent raters (MD, MX, FT, ND). Raters were blinded to one another’s assessments, and any disagreements were resolved by a third reviewer (CZ) to ensure consistency and adherence to the eligibility criteria. Subsequently, the full-text of the remaining entries was reviewed once. To ensure quality control and reduce intrapersonal variability, one-third of the full-text reviews were independently checked by an additional rater. Reasons for exclusion were documented at the title, abstract and full-text stage.

**Fig. 1.**
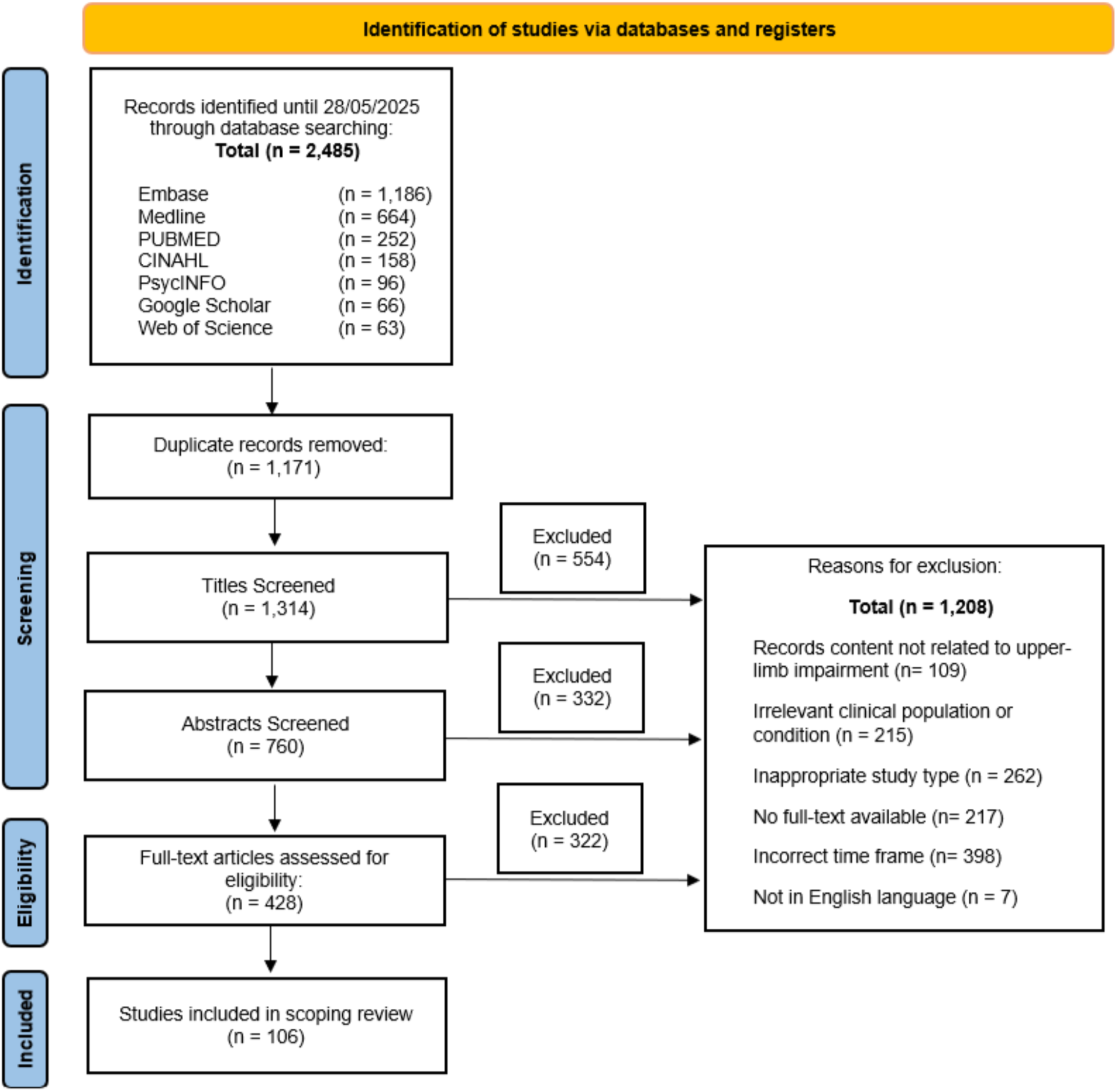
Flow diagram illustrating numbers of articles identified, screened, assessed for eligibility, and included in the review.

### 2.4. Data Charting Process

The extraction criteria were established a priori. The following information was extracted: title, author(s), year of publication, study design, sample demographics (including sample size, age, gender distribution), country of origin of the study, and time post-stroke. Time post-stroke data were standardized and converted into days for consistency (e.g., 1 month = 30 days). Regarding UL outcome measures, we focused on clinical assessments spanning motor, sensory, and global measures of UL capacity, as well as measures of brain structure and function. Details on follow-up assessments, including the nature and timing of these evaluations, were also systematically recorded. A complete list of the extracted data elements is provided in the supplementary material **(Suppl. Material)**.

The primary focus of this scoping review was to identify which UL assessments are employed in stroke research. Accordingly, the data extraction process was focused on collating information on the types and frequencies of assessments employed. It is important to note that this review does not extract data on study findings, nor does it include any formal assessment of study quality.

## 3. Results

### 3.1 Search results

The initial search yielded a total of 2,485 records, from which 1,171 duplicates were removed using EndNote 20 (Clarivate Analytics, PA, USA) and thorough manual verification. Following the application of predefined inclusion and exclusion criteria, an additional 1,208 records were excluded during the screening process. This resulted in a final set of 106 original studies deemed eligible for data extraction and synthesis. These publications serve as the foundation of our analysis of UL assessments in acute stroke research. The detailed article selection process is outlined in **Fig. 1**. A complete list of the included publications is provided in the supplementary material **(Suppl. Material)**.

### 3.2 Discrepancies in the definition of ‘acute’ stroke

Clear definitions are essential for accurate assessment timing in acute stroke research. The widely accepted temporal definition of ‘acute’ stroke refers to the first 7 days post-stroke onset, meaning the assessments evaluating acute UL capacity should occur within this timeframe (Bernhardt et al., 2017). However, we identified notable inconsistencies among the included studies. Specifically, 39 papers labelled as ‘acute’ conducted initial assessments beyond this 7-day window, effectively diverging into the early subacute phase. Additionally, 4 studies failed to report time post-stroke at their initial UL assessments despite categorising their cohorts as acute stroke survivors.

### 3.3 Study Demographics

The highest number of publications originated from the United States of America (n=16) and the China (n=16, **Fig. 2a**). Roughly half of the studies were observational in design, with 37% being longitudinal, 12% cross-sectional, and 3% case studies. Interventional studies accounted for the remaining half of the studies, comprising 26% clinical trials and 22% prospective cohort studies (**Fig. 2b**). Sample sizes ranged from 1 to 1471 participants (*M* = 69.8, *Med* = 40.0, *STD* = 154.9, *SE* = 15.0), with 33.0% of studies having 25 or fewer subjects (**Fig. 2c**). On average more male than female participants were included in these studies (Male: *M* = 40.6, *Med* = 21.0, *STD* = 88.2, *SE* = 8.6; Female: *M* = 29.5, *Med* = 18, *STD* = 67.3, *SE* = 6.5; *t (104)* = 4.48, *p* <0.001). The mean age across studies was 64.0 years (*Med* = 64.8, *STD* = 8.9, *SE* = 0.9, **Fig. 2d**). Amongst the studies providing an age range (55.7% of the studies) only 33.9% have a minimum age below 35 years, illustrating the lack of research in younger stroke survivors (**Fig. 2d**).

**Fig. 2.**
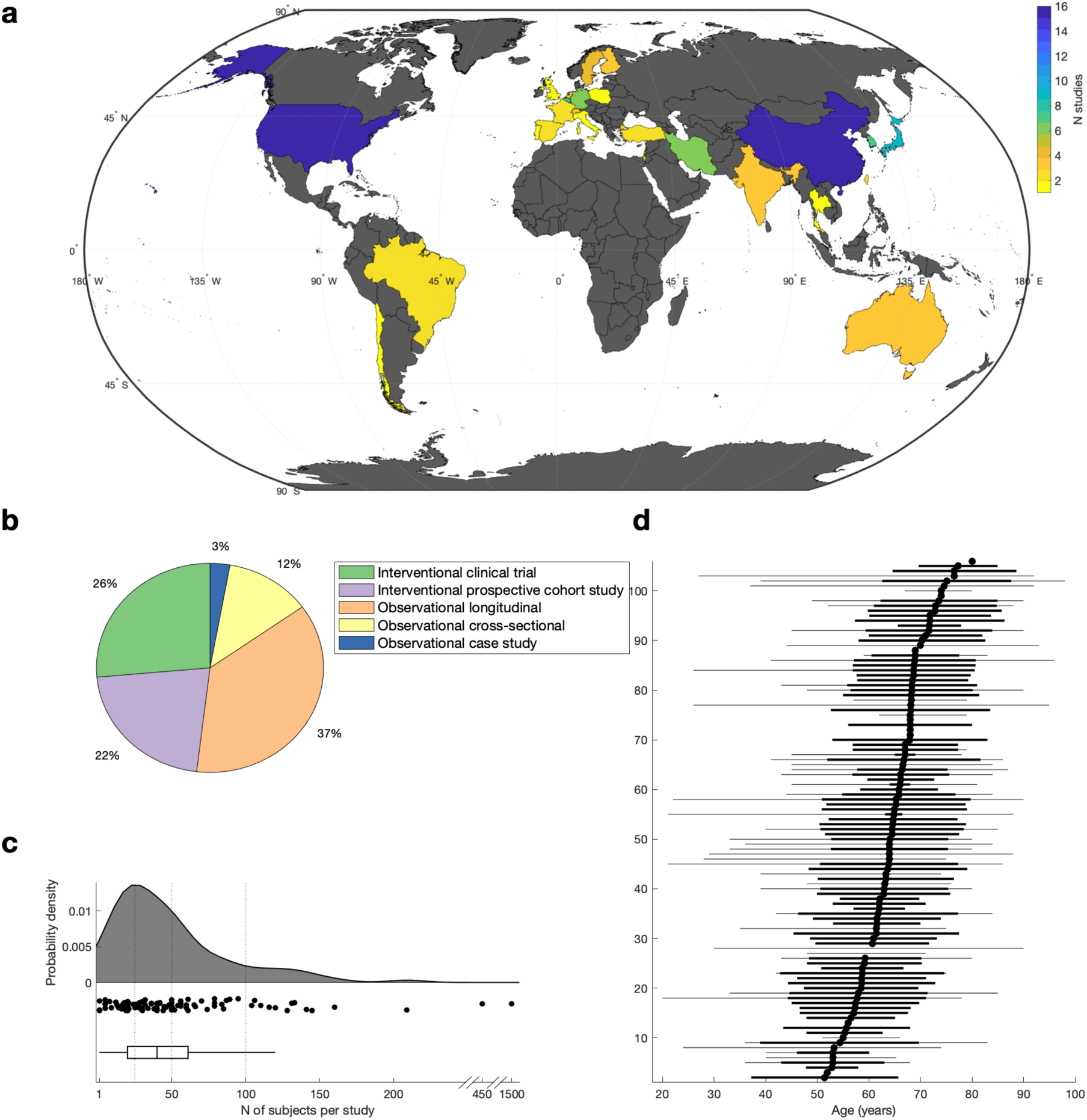
Study Demographics. a) Geographical distribution of included studies. b) Included studies categorised by study type. c) Number of subjects per study. Shown is the probability function (top), the corresponding individual datapoints (middle), and the boxplot (bottom). Horizontal line indicates N = 25, N = 50, and N = 100. d) Age of subjects per study. Each row represents one study in the scoping review, which are sorted based on the average age (i.e., mean or median, depending on data availability). Average age is shown as a block dot, variance (i.e., standard deviation) is shown as thick black line, and age range is shown as thin grey line.

### 3.4 Clinical global assessment after stroke

At least one clinical measure of global capacity was used in 76.4% of the studies. The National Institutes for Health Stroke Scale (NIHSS) was the most frequently employed measure, appearing in 58.5% of articles. This scale is renowned for its robustness in evaluating stroke severity in acute settings (Chalos et al., 2020; Lyden, 2017). We found that 55 studies used the NIHSS exclusively as a measure to determine stroke severity at baseline, aligning with the SRRR recommendations for routinely early assessment (Kwakkel et al., 2017). Meanwhile, 17 studies used the NIHSS scale as an outcome measure to track changes over time. This dual application highlights the lack of consensus on the most appropriate use of the NIHSS.

Following the NIHSS, the modified Rankin Scale (mRS) was the second most prevalent assessment tool, appearing in 28.3% of studies. Additionally, the Barthel Index (BI), which evaluates the ability to perform essential activities of daily living (ADLs), was employed in 24.5% of studies. The prominence of these tools suggests that stroke researchers prioritise both stroke severity and functional independence as key metrics for assessing overall patient outcomes.

Less commonly used measures include the Stroke Impact Scale (SIS), and Functional Independence Measure (FIM), featuring 6.6% and 5.7% of studies. Additionally, we identified four other global measures, each appearing in 2% or less of the included articles, as illustrated in **Fig. 3a**. These findings collectively emphasise a preference for established, validated global assessment tools within the field.

**Fig. 3.**
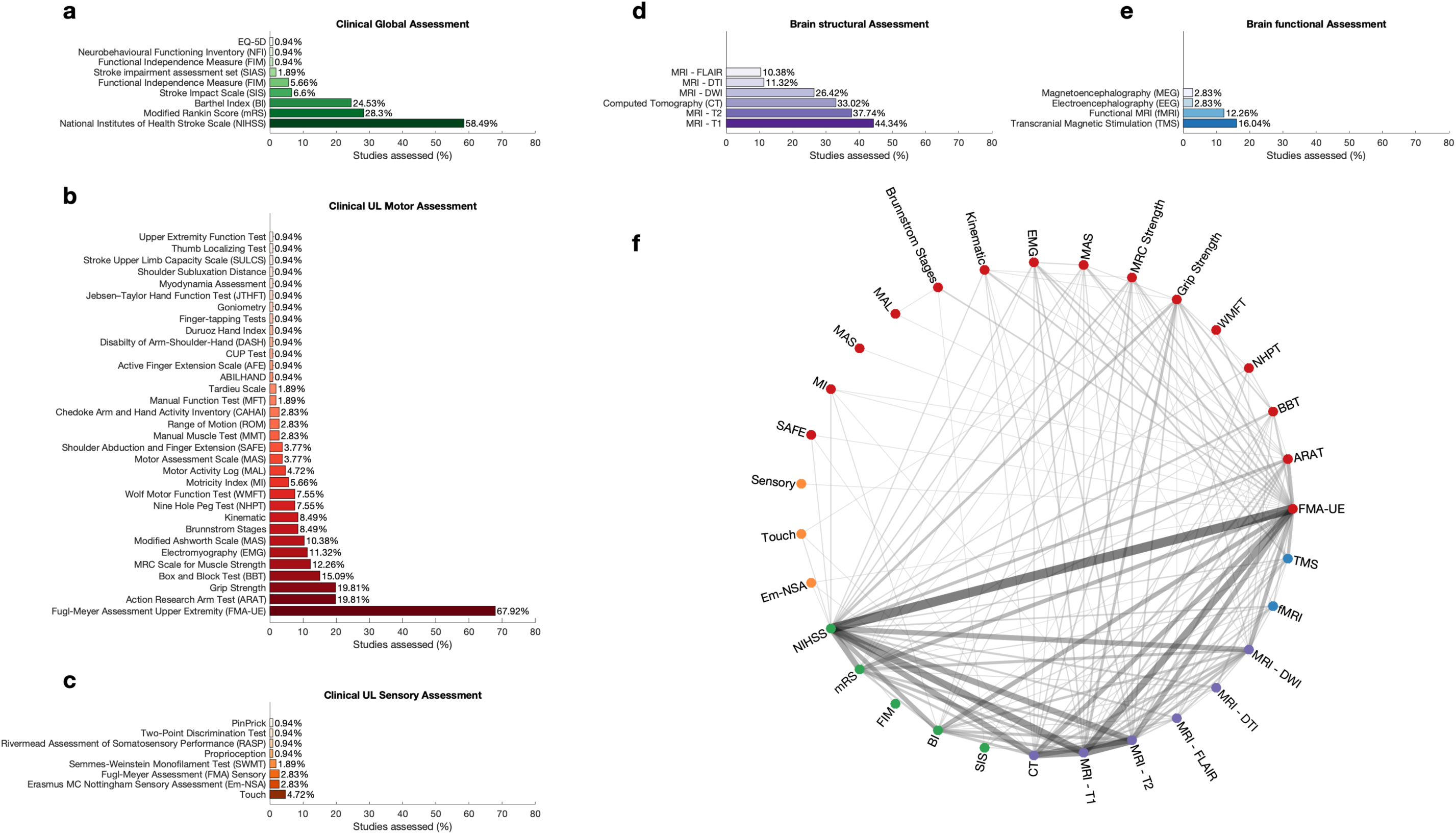
Percentage of Assessment categorised by Assessment type. a) Clinical Global Assessment. Assessments are ranked and coloured by the frequency of which they are used. b) Clinical UL Motor Assessment. Assessments are ranked and coloured by the frequency of which they are used. c) Clinical UL Sensory Assessment. Assessments are ranked and coloured by the frequency of which they are used. d) Brain structural Assessment. Assessments are ranked and coloured by the frequency of which they are used. e) Brain functional Assessment. Assessments are ranked and coloured by the frequency of which they are used. f) Connectivity plot indicating which Assessments are obtained together in one study. The colour of each node corresponds to the assessment category; green for global clinical assessments, red for motor assessments, orange for sensory assessments, blue for measures of brain function, and purple for measures of brain structures. Colour and thickness of the edge between two nodes indicates the frequency of which two assessments are used together in one study. Connections that occur less than 1% are omitted from the plot.

### 3.5 Clinical upper-limb sensorimotor assessments after stroke

In total, 41 distinct clinical outcome measures were identified, spanning both sensory and motor assessments.

#### Motor assessment

95.3% of the studies used at least one clinical measure of UL motor assessment. However, the landscape of UL motor assessments used was notably heterogeneous (**Fig. 3b**). The Fugl-Meyer Upper Extremity Assessment (FMA-UE) emerged as the predominant tool (67.9%). Grip Strength, typically assessed using a dynamometer, and the Action Research Arm Test (ARAT), were each reported in 19.8% of the studies. The Box and Block Test (BBT) appeared in 15.1% of studies. Beyond these leading assessments, our search uncovered an additional 29 outcome measures, of which 18 were reported in 3% or less of studies.

#### Sensory assessment

In comparison, only 14.2% of the studies used at least one measure of UL sensory assessment. Amongst these (**Fig. 3c**), touch was most used (4.7%), followed by the Erasmus MC Nottingham Sensory Assessment (Em-NSA) and the Fugl-Meyer Upper Extremity Sensory subscale (FMA-S), featuring each in about 2.8% of articles. The Semmes-Weinstein Monofilament Test (SWMFT) accounted for 1.9% of studies. Other outcome measures were employed even less frequently. The scarcity of sensory assessments highlights a gap, as sensory and motor capacity can interact.

This broad set of tools reflects the complexity and variability of measuring UL capacity in acute stroke research.

### 3.6 Brain structural assessments after stroke

Structural neuroimaging provides a unique window into stroke-related changes in both grey and white matter. Our findings showed that 62.3% of the studies employed some form of brain structural assessment, with Magnetic Resonance Imaging (MRI) being the most routinely employed technique (**Fig. 3d**). T1-weighted MRI was the most frequently reported imaging modality (44.3%), followed by T2-weight MRI (37.7%), and Computed Tomography (CT, 33.0%). Diffusion Weighted Imaging (DWI) was used in 26.4%, while Diffusion Tensor Imaging (DTI) and Fluid-Attenuated Inversion Recovery (FLAIR) occurred in 11.3% and 10.4% of the studies. Among the studies employing structural neuroimaging, a little under half (45.5%) applied it exclusively for stroke diagnosis confirmation, without using it for further outcome evaluation.

### 3.7 Brain functional assessments after stroke

Measures of brain function offer valuable insights into stroke-related changes in brain function (e.g., functional connectivity, cortical excitability). However, our findings indicate a comparatively lower use of functional assessments, with only 29.2% of studies (**Fig. 3e**). Transcranial Magnetic Stimulation (TMS) was the most often used technique, seen in 16.0% of articles, with motor evoked potentials (15.1%) and motor threshold (10.4%) being most frequently assessed. More complex measures, such as recruitment curve, short-interval intracortical inhibition (SICI), long-interval intracortical inhibition (LICI), and intracortical facilitation (ICF), were not assessed in any of the studies. Functional MRI (fMRI) was used in 12.3% of articles, while Electroencephalography (EEG) and Magnetoencephalography (MEG), were only used each in 2.8% of studies. Other measures such as Functional near-infrared spectroscopy (fNIRS) or positron emission tomography (PET) were not used at all. Amongst fMRI, EEG, and MEG studies, resting state was collected more often than task data. If task data were collected, the tasks differed across studies including peripheral nerve stimulation, wrist extension, finger tapping, and active or passive movement of the index finger.

### 3.8 Which assessments are conducted together

Next, we asked which assessments tools were commonly performed together (**Fig. 3f**). FMA-UE was often performed with a clinical global assessment tool, most commonly the NIHSS, but also the mRS and BI. Structural imaging was often combined with both the FMA-UE and the NIHSS, and MRI T1-weighted, MRI T2-weighted, and CT were often conducted in the same study.

### 3.9 Follow-up Assessment

84.9% of the studies conducted at least one follow-up assessment. Figure 4 depicts the distribution of clinical follow-up assessments across studies, measured at different intervals post-baseline. Despite substantial variability regarding the timing of the follow-up assessments (range = 1 day to 4 years), 81.1% of studies performed their initial follow-up assessment within 3 months post-baseline. Amongst the subset of clinical trials, only 59.3% conducted a follow-up at 3 months, which is strongly recommended (Kwakkel et al., 2017). Following the 3-month mark, the frequency of follow-up assessments declines markedly, with only 10.4% of studies conducting follow-ups beyond 6 months.

**Fig. 4.**
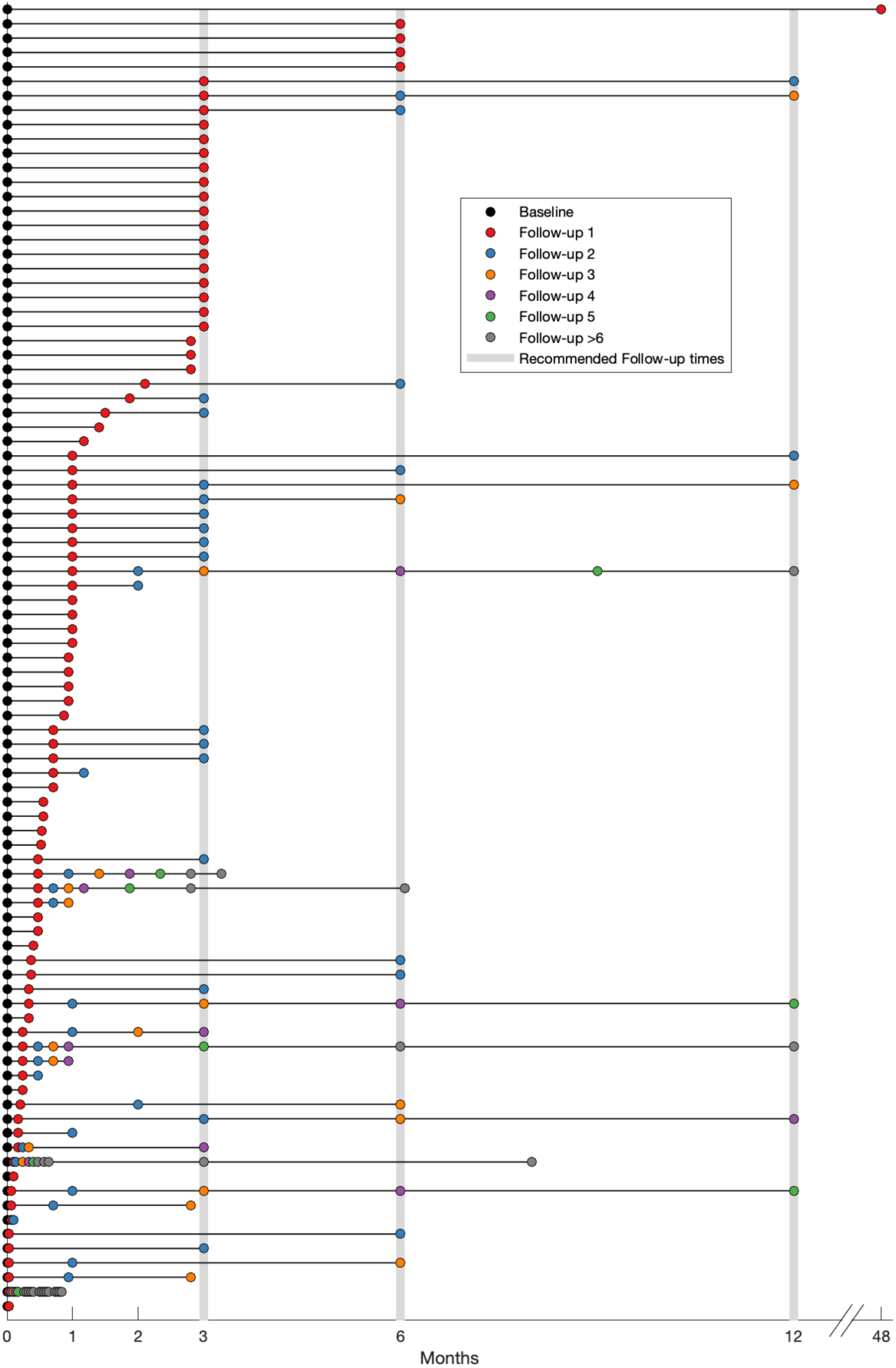
Timing of follow-up assessments. Each row represents one study in the scoping review, which are sorted based on the timing of the first Follow-up. The Baseline assessment is highlighted in black. Follow-up assessments 1-5 are represented by coloured circles, Follow-up assessments larger than 5 are represented as grey circles. Grey vertical lines indicate the recommended time points for follow-up assessments.

## 4. Discussion

This scoping review aimed to evaluate how current research practices in acute stroke are aligned with the SRRR recommendations, specifically regarding the assessment of UL sensorimotor capacity. Our findings revealed progress in some domains, particularly global clinical assessments, but also highlighted substantial inconsistencies, gaps, and underutilised opportunities for integrating multimodal data.

### 4.1 Why SRRR? A Framework for Consistency

The SRRR provides a research-driven framework for stroke recovery research, offering a valuable benchmark for standardisation in measurement timing, tool selection, and multimodal assessment (Kwakkel et al., 2017). Its structure facilitates data comparability, supports meta-analyses, and encourages international collaboration. Our study evaluates the degree to which acute stroke studies adhere to SRRR-specified measures and timelines.

### 4.2 Clinical Assessments: Global and Upper Limb

#### Global Assessments: Regular but Inconsistently Applied

There was a largely consistent use of global clinical measures, particularly the NIHSS, employed in 58% of studies. The NIHSS remains a foundational tool for assessing baseline stroke severity; however, its dual application, as both a severity marker and an outcome measure, reflects a misinterpretation of its intended purpose (Kwakkel et al., 2017).

Other global tools such as the mRS and BI were also frequently used, often to assess disability or functional independence. These choices suggest a research focus on broad outcomes.

#### Upper Limb Motor Assessments: Dominated by FMA-UE

In line with SRRR guidelines, the FMA-UE was the most frequently used UL motor outcome. Its robust validation and sensitivity to impairment-level changes justify its prevalence. In contrast, only ∼20% of studies employed the ARAT, which evaluates activity limitations, a core domain under the ICF model (World Health Organization, 2001). This substantial gap between impairment and activity assessments reveals an overemphasis on body structure/function and an underutilisation of tools that assess functional application in daily life.

Furthermore, the identification of 41 different UL clinical measures across studies highlights the considerable heterogeneity within the field. While this diversity reflects attempts to capture different dimensions of recovery, it also complicates efforts to synthesise findings across studies. Greater alignment on core assessment tools, including validated measures of both impairment and activity, is needed to improve cross-study comparability.

#### Sensory and Kinematic Assessments: A Persistent Oversight

Despite sensory deficits being common and influential in shaping sensorimotor recovery trajectories, sensory capacity was assessed in only 14% of studies. The relative neglect of the sensory system is notable, especially since deficits in touch, proprioception, and sensory integration can profoundly affect motor execution and recovery. Tools such as the Em-NSA, SWMFT, or FMA sensory subscales were rarely used. This gap calls for greater attention to sensory deficits in future acute stroke studies, both for more comprehensive patient profiling and for developing sensory biomarkers - identified by the SRRR as a future research priority (Boyd et al., 2017)

Additionally, the SRRR emphasises the use of kinematic measures to complement clinical tools. These assessments, such as 2D or 3D reaching tasks, can differentiate between behavioural restitution and compensation strategies (Kwakkel et al., 2019). Yet their presence in acute stroke research remains limited, representing another opportunity for methodological expansion.

### 4.3 Brain Imaging: Diagnostic Use Over Biomarker Development

Structural neuroimaging was present in 62% of studies, but roughly half used it solely for diagnostic confirmation rather than for outcome tracking or biomarker identification. While T1- and T2-weighted MRI, CT, and DWI were common, more advanced measures like DTI were underused. This suggests untapped potential for using structural data, not just for inclusion criteria, but also to inform prognosis and guide treatment.

Functional assessments were even less prevalent. Techniques such as TMS and fMRI were used in 16% and 12% of studies, respectively. These low numbers may reflect cost, logistical complexity, or variability in interpretation, but they also limit insights into brain reorganisation post-stroke. Notably, advanced TMS paradigms like SICI and ICF were absent, and functional modalities such as PET or fNIRS were not used at all. To align with SRRR priorities, greater adoption of these tools, especially in multimodal formats, is needed to establish robust, reproducible biomarkers.

### 4.4 Joint Assessments: Fragmented Methodologies

Our data suggest that only a limited number of studies incorporated multiple modalities (e.g., combining FMA-UE, NIHSS, and neuroimaging). The lack of integrated assessments undermines the ability to understand the interaction between brain structure, brain function, and motor capacity. Future research should focus on multimodal designs that simultaneously evaluate clinical, structural, and functional metrics to develop a more holistic understanding of post-stroke recovery.

### 4.5 Follow-up Assessments: Variability and Gaps

While ∼85% of studies included follow-up assessments, the timing varied widely. Although ∼82% conducted follow-up within 3 months, only just over half of clinical trials included the SRRR-mandated 3-month follow-up. Few studies included 6- or 12-month assessments, despite their importance for tracking longer-term outcomes. This variability may stem from logistical challenges, funding limitations, and participant retention concerns, but it introduces considerable noise into longitudinal comparisons.

Furthermore, imprecise reporting of time points - such as using ‘1 month’, ‘30 days’, or ‘4 weeks’ interchangeably - introduces ambiguity. Even small timing differences can bias interpretation, especially in the early recovery period, where functional gains may occur rapidly.

### 4.6 Demographic and Reporting Inconsistencies

We observed a systematic underreporting of demographic details. While mean and median ages were often provided, granular information about age distribution within cohorts was missing. Only 34% of studies included participants under age 35, and males were significantly overrepresented. This is concerning given epidemiological evidence suggesting that while stroke incidence is higher in men, women may experience more severe outcomes and have different recovery trajectories (Hiraga, 2017; Reeves et al., 2008). The lack of gender-balanced samples raises questions about the generalisability of current findings and suggests a potential research bias.

In addition, inconsistent definitions of the acute phase, despite SRRR defining it as 1-7 days post-onset, persist across studies and even between countries. Greater global harmonisation of terminology and measurement timing is needed to support robust data synthesis.

### 4.7 Conclusion

Our scoping review highlights both strengths and limitations in the current landscape of acute stroke UL assessment. While tools like the NIHSS and FMA-UE are widely used, inconsistency in application, underuse of sensory and activity-based assessments, limited adoption of functional imaging, and variability in follow-up protocols all undermine data comparability. To advance the field, future research must embrace more standardised, multimodal, and demographically inclusive approaches, in full alignment with SRRR guidelines.

## Data Availability

NA - scoping review

## Supplementary Material

**Supplementary Table 1.** Search strategies limited to publications from 01.08.2017 - 28.05.2025.

| Database | Platform | Search string |
| --- | --- | --- |
| **Embase** | Ovid | *("arm":ti,ab,kw OR "upper limb":ti,ab,kw OR "upper extremity":ti,ab,kw) AND ("movement":ti,ab,kw OR "motor":ti,ab,kw) AND ("acute":ti,ab,kw OR "early":ti,ab,kw) AND "stroke":ti,ab,kw* |
| **MEDLINE** | Ovid | *("arm":ti,ab,kw OR "upper limb":ti,ab,kw OR "upper extremity":ti,ab,kw) AND ("movement":ti,ab,kw OR "motor":ti,ab,kw) AND ("acute":ti,ab,kw OR "early":ti,ab,kw) AND "stroke":ti,ab,kw* |
| **PubMed** | PubMed | *((("arm"[Title/Abstract] OR "upper limb"[Title/Abstract] OR "upper extremity"[Title/Abstract]) AND "acute"[Title/Abstract] AND "stroke"[Title/Abstract] AND "motor"[Title/Abstract]) NOT "sub-acute"[Title/Abstract]) NOT "subacute"[Title/Abstract])* |
| **CINAHL** | EBSCO | *("arm":AB OR "upper limb":AB OR "upper extremity":AB) AND ("motor":AB OR "movement":AB) AND ("acute":AB AND "stroke":AB NOT "subacute":AB)* |
| **PsycINFO** | Ovid | *("arm":ti,ab OR "upper limb":ti,ab OR "upper extremity":ti,ab) AND ("movement":ti,ab OR "motor":ti,ab) AND ("acute":ti,ab OR "early":ti,ab) AND "stroke":ti,ab* |
| **Google Scholar** | Google Scholar | *allintitle: acute stroke motor arm OR limb OR extremity –subacute* |
| **Web of Science** | Web of Science | *(((((TI=(motor)) OR TI=(movement) AND TI=(arm) OR TI=(upper limb) OR TI=(upper extremity) AND TI=(stroke) AND TI=(acute)* |

### Comprehensive list of data extraction elements

- **Paper Specifics**

∘ Author
∘ Year
∘ Title
∘ Journal
∘ URL (DOI or stable link)
- **Study Type**

∘ Interventional

▪ Yes/No
▪ Clinical trial (1/0)
▪ Clinical trial phase
▪ Clinical trial link
∘ Observational

▪ Longitudinal
▪ Cross-sectional
▪ Case report(s)
▪ Other (specify)
- **Demographics**

∘ Number of sites
∘ Recruitment period
∘ Sample size (N)
∘ Male (n)
∘ Female (n)
∘ Age (mean, range)
∘ Time post-stroke at inclusion
∘ City
∘ Country
∘ Notes (e.g., median age, key cohort characteristics)
- **Clinical Assessments (1 = included, 0 = not included)**

∘ **Upper Limb Motor**

▪ Fugl-Meyer Assessment Upper Extremity (FMA-UE)
▪ Action Research Arm Test (ARAT)
▪ Box and Block Test (BBT)
▪ Nine Hole Peg Test (NHPT)
▪ Wolf Motor Function Test (WMFT)
▪ Grip Strength
▪ Medical Research Council (MRC) Scale
▪ Lovett Scale
▪ Ashworth Scale
▪ Electromyography (EMG)
▪ Chedoke Arm and Hand Activity Inventory (CAHAI)
▪ Kinematic assessments
- ∘ **Upper Limb Sensation**

▪ PinPrick
▪ Fugl-Meyer Sensory
▪ Touch
- ∘ **Global**

▪ National Institutes of Health Stroke Scale (NIHSS)
▪ Modified Rankin Scale (mRS)
▪ Functional Independence Measure (FIM)
▪ 36-item Short-Form Health Survey (SF-36)
▪ EQ-5D
▪ Barthel Index (BI)
▪ Stroke Impact Scale (SIS)
- ∘ Notes (e.g. other tests, used for inclusion/exclusion criteria)
- **Clinical Assessments Follow-Up**

∘ Yes/No
∘ When
∘ What
- **Brain Structure (1/0)**

∘ Computed Tomography (CT)
∘ MRI T1
∘ MRI T2
∘ MRI FLAIR
∘ MRI DTI
∘ MRI DWI
∘ MRI Proton
∘ MRI GRASE
∘ Other (specify)
∘ Notes
- **Brain Structure Follow-up**

∘ Yes/No
∘ When
∘ What
- **Brain Function (1/0)**

∘ Electroencephalography (EEG)

▪ Rest
▪ Task
▪ Task category
∘ Magnetoencephalography (MEG)

▪ Rest
▪ Task
▪ Task category
∘ Functional MRI (fMRI)

▪ Rest
▪ Task
▪ Task category
∘ Functional Near-Infrared Spectroscopy (fNIRS)

▪ Rest
▪ Task
▪ Task category
∘ Positron Emission Tomography (PET)

▪ Rest
▪ Task
▪ Task category
∘ Transcranial Magnetic Stimulation (TMS)

▪ MEP
▪ Motor Threshold
▪ Recruitment curve
▪ Short-Interval Intracortical Inhibition (SICI)
▪ Silent Period (SP)
▪ Long-Interval Intracortical Inhibition (LICI)
▪ Intracortical Facilitation (ICF)
∘ Other (specify)
∘ Notes
- **Brain Function Follow-Up**

∘ Yes/No
∘ When
∘ What
- **Other Biological Measures (1/0)**

∘ Blood biomarkers
∘ Genetic testing
∘ Other (specify)

